# How strong is the epidemiological evidence to support any potential protective role for vitamin D levels on COVID-19 infections and mortality? - A time-series analysis of European Populations

**DOI:** 10.1101/2020.11.20.20235705

**Authors:** Samer Singh

## Abstract

A potential protective role of vitamin D serum levels on overall adverse outcomes of SARS-CoV-2 infection or COVID-19 on populations had been suggested previously based upon single-point cross-sectional analysis of 8 April 2020 data from 20 European countries assuming comparable underlying confounding variables for these populations, at an early stage of the current pandemic. Comparative time-series cross-sectional analysis of the COVID-19 data from 12 March (early pre-peak) to 26 July (late post-peak of infections) 2020 was performed to assess the strength of the assertion. The study subjects included 1,829,634 COVID-19 cases (11.11% of total worldwide) and 179,135 associated deaths (27.45 % of total worldwide) on 26 July 2012. Previously suggested cross-sectional study design and methodology could not consistently and significantly (p-value≥0.05) support the notion of the potential protective role of the mean serum vitamin D levels of the populations on COVID-19 incidence and mortality. However, the exponential correlative model, as well as alternative simple regression analysis on *ln* and *Log*_*10*_ transformed COVID-19 data for the time period indicated improved consistently negative covariation with vitamin D levels. Additionally, the later methodology increased the predictive potential for explaining the variability in data [*R*^*2*^ by 1.27-1.96 fold, adjusted-*R*^*2*^ by 1.33-2.47, *p-*value=0.0457-0.0035, for cases/million; *R*^*2*^ by 1.81-2.67, adjusted-R^2^ by 2.21-3.74 fold for deaths/million, *p-*value=0.0049-0.0228). Considering, the established role of vitamin D in immune system functioning randomized well-controlled trials may be suggested to evaluate/assess the potential protective role of vitamin D in reducing the COVID-19 impact on populations.

## INTRODUCTION

The urgency of the COVID-19 pandemic caused by SARS-CoV-2 had suddenly posed a great majority of researches to focus on understanding COVID-19 and explore ways to mitigate its effects on the worldwide human population. By 26 July 2020, more than 16.4 million cases of COVID-19 had been reported with more than 652 thousand lives lost [1]. European countries (including Turkey) had been disproportionately affected by COVID-19, accounting for 18.34% cases and 31.73% of deaths. Many potential protective variables for the populations have been suggested based upon statistical analyses, e.g. Trained immunity, BCG vaccination coverage/policy, Vitamin D levels, Zinc levels, *etc* [2-6 and References therein]. Vitamin D serum levels in European countries had been suggested to be potentially playing a protective role based upon a one point cross-sectional analysis of the data by Ilie et al. [5]. However, cautionary notes by us [7] and more recently by Maruotti *et al*. [8] have been raised on the methodology and analysis. The letter by Maruotti et al. beautifully articulates the existing general fascination of interpreting any exploratory correlation as to have a cause and effect relationship by people at large. From time to time various scientists and bodies keep coming forward to educate the general audience on the same [References in 8]. In the light of our suggestions and those made by Maruotti et al., we explore the potential evidence for the protective role of populations mean vitamin D levels on COVID-19 incidence and mortality and present a detailed time-series analysis of the COVID-19 data of the same twenty European countries whose mean serum vitamin D levels are known and had been analyzed previously.

We hypothesized that if indeed vitamin D levels may have any potential protective role in COVID-19 as suggested previously, the COVID-19 impact on the select European countries with different vitamin D levels would consistently negatively covary/correlate rather than appearing as an effect of random fluctuation or seasonality just observable on 8 April 2020. The methodology previously employed failed to consistently support the potential protective role of vitamin D over the time period [5]. However, the methodology suggested by us [7] consistently supported the potential protective role of vitamin D levels on COVID-19 incidences and mortality during the study period 26 March 2020 to 26 July 2020.

## MATERIALS AND METHODS

Twenty European countries for which average vitamin D levels of the populations are reported in the literature [5], have comparable exposure to UV rays but have displayed variable COVID-19 impact, and have been analyzed in the recent past for assessing covariation of the Vitamin D levels in the populations with the overall COVID-19 impact on them [5,7] were selected for the current analysis. The time-series data of total COVID-19 cases and deaths reported per million populations for the time period starting from 12 March to 26 July 2020 (Supplementary Table 1) were taken from a previous publication [5] and the coronavirus pandemic data portal https://www.worldometers.info/coronavirus/ [1]. The country-specific mean serum vitamin D levels (nmol/L) data were taken from the previous publication [5]. The study subjects included a total of 1,829,634 cases and 179135 deaths translating into 11.11% cases and 27.45% deaths from COVID-19 worldwide. Data transformation (*log* and *ln*) and basic statistics calculations (Pearson correlation coefficient, Regression analysis, curve-fitting - calculation of best-fit trend line) were performed using Microsoft Excel as done previously [6,7].

## RESULTS

The simple linear regression analysis previously used to propose potential protective covariation of the mean Vitamin D levels of the populations with COVID-19 impact based upon a single day data was found inadequate to support such an assertion when retested and applied over a period of time at a commonly used and previously used significance p-value cutoff (<0.05) (Table 1 and Fig. 1 A; See supplementary file 1 for COVID-19 incidence and mortality data along with estimates of vitamin D for the population, basic statistical estimates, and the regression analysis). The negative correlation between vitamin D levels and COVID-19 infections observed in per million population was found to be statistically significant (p-value <0.05) only between 12 April to 12 June 2020, and remained non-significant thereafter upto the end of the current analysis period, i.e., 26 July 2020. The correlation between deaths per million population with vitamin D levels never reached the suggested statistical significance level (p-value <0.05) during the whole study period, *i*.*e*., 8 April to 26 July 2020, (*p-value* for correlation varied from 0.0535 to 0.1550). The presence of heteroscedasticity in the early infection data (data not shown) further makes the regression analysis prone to over/under-estimation.

**Table 1.** Time-series correlation and regression analysis of vitamin D mean serum level (nmol/L) and COVID-19 incidence (top panel) and mortality per million (bottom panel) data of European countries. *Note* the estimates of correlation *r*(20)/R-value, R^2^, adjusted-R^2^, the *p*-value for the exponential model, and the linear regression of *ln* transformed data will be same as that displayed for *log*_*10*_ transformed data but they would not suppress the apparent variability in data allowing easy variable discovery (compare Fig. 1D with 1C and 1B, see discussion). The highlighted 8 April 2020 data was previously analyzed in a cross-sectional study [5].

|  |  | COVID-19 CASES per MILLION (CpM) vs POPULATIONS' Vit. D LEVELS (nmol/L) |  |  |  |  |  |  |  |  |  |  |  |
| --- | --- | --- | --- | --- | --- | --- | --- | --- | --- | --- | --- | --- | --- |
|  |  | Data-Date | 12 Mar | 26 Mar | 8 Apr | 12 Apr | 26 Apr | 12 May | 26 May | 12 Jun | 26 Jun | 12 Jul | 26 Jul |
| Simple linear Regression | Corr r(20)/R value |  | -0.2760 | -0.4080 | -0.4435 | -0.5072 | -0.5448 | -0.5412 | -0.5265 | -0.4727 | -0.4054 | -0.3908 | -0.4034 |
|  | R Square |  | 0.0762 | 0.1665 | 0.1967 | 0.2572 | 0.2968 | 0.2929 | 0.2772 | 0.2235 | 0.1644 | 0.1527 | 0.1627 |
|  | Adjusted R Square |  | 0.0248 | 0.1202 | 0.1521 | 0.2160 | 0.2577 | 0.2536 | 0.2370 | 0.1803 | 0.1180 | 0.1057 | 0.1162 |
|  | Standard Error |  | 159.2335 | 755.9334 | 1040.5152 | 1170.7413 | 1280.8313 | 1361.5566 | 1426.5450 | 1557.9696 | 1738.0987 | 1844.5105 | 1915.5266 |
|  | F-value |  | 1.4837 | 3.5957 | 4.4079 | 6.2335 | 7.5960 | 7.4557 | 6.9031 | 5.1799 | 3.5411 | 3.2446 | 3.4985 |
|  | p-value |  | 0.2389 | 0.0741 | 0.0501 | 0.0225 | 0.0130 | 0.0137 | 0.0171 | 0.0353 | 0.0761 | 0.0884 | 0.0778 |
|  |  | LOG TRANSFORMED (LOG<sub>10</sub>) CASES per MILLION (CpM) vs POPULATIONS' Vit. D LEVELS (nmol/L) |  |  |  |  |  |  |  |  |  |  |  |
|  |  |  | 12 Mar | 26 Mar | 8 Apr | 12 Apr | 26 Apr | 12 May | 26 May | 12 Jun | 26 Jun | 12 Jul | 26 Jul |
| Exponential Model<br>or Log<sub>10</sub><br>Transformed<br>Variable- Linear<br>Regression | Corr r(20)/R value |  | -0.0526 | -0.4515 | -0.5685 | -0.6205 | -0.6202 | -0.6101 | -0.6015 | -0.5786 | -0.5544 | -0.5476 | -0.5477 |
|  | R-square |  | 0.0028 | 0.2039 | 0.3232 | 0.3850 | 0.3846 | 0.3722 | 0.3618 | 0.3348 | 0.3074 | 0.2999 | 0.3000 |
|  | Adjusted R Square |  | -0.0526 | 0.1597 | 0.2856 | 0.3508 | 0.3505 | 0.3373 | 0.3263 | 0.2978 | 0.2689 | 0.2610 | 0.2611 |
|  | Standard Error |  | 1.0500 | 0.4996 | 0.3591 | 0.3320 | 0.2983 | 0.2951 | 0.2952 | 0.3035 | 0.3116 | 0.3081 | 0.3030 |
|  | F-value |  | 0.0499 | 4.6101 | 8.5972 | 11.2685 | 11.2514 | 10.6713 | 10.2045 | 9.0578 | 7.9891 | 7.7107 | 7.7129 |
|  | p-value |  | 0.8258 | 0.0457 | 0.0089 | 0.0035 | 0.0035 | 0.0043 | 0.0050 | 0.0075 | 0.0112 | 0.0124 | 0.0124 |
|  |  | DEATHS per MILLION (DpM) vs POPULATIONS' Vit. D LEVELS (nmol/L) |  |  |  |  |  |  |  |  |  |  |  |
|  |  | Data-Date | 12 Mar | 26 Mar | 8 Apr | 12 Apr | 26 Apr | 12 May | 26 May | 12 Jun | 26 Jun | 12 Jul | 26 Jul |
| Simple linear Regression Analysis | Corr r(20)/R value |  | -0.1948 | -0.3683 | -0.4378 | -0.4193 | -0.3990 | -0.3745 | -0.3596 | -0.3448 | -0.3340 | -0.3303 | -0.3291 |
|  | R Square |  | 0.0379 | 0.1357 | 0.1917 | 0.1758 | 0.1592 | 0.1403 | 0.1293 | 0.1189 | 0.1116 | 0.1091 | 0.1083 |
|  | Adjusted R Square |  | -0.0155 | 0.0876 | 0.1468 | 0.1300 | 0.1125 | 0.0925 | 0.0809 | 0.0699 | 0.0622 | 0.0596 | 0.0588 |
|  | Standard Error |  | 3.7722 | 33.3574 | 87.3958 | 115.6291 | 175.3307 | 213.2015 | 229.3434 | 241.0568 | 246.1939 | 248.9276 | 250.2122 |
|  | F-value |  | 0.7099 | 2.8252 | 4.2684 | 3.8389 | 3.4087 | 2.9372 | 2.6728 | 2.4285 | 2.2609 | 2.2037 | 2.1864 |
|  | p-value |  | 0.4105 | 0.1101 | 0.0535 | 0.0658 | 0.0814 | 0.1037 | 0.1194 | 0.1366 | 0.1500 | 0.1550 | 0.1565 |
|  |  | LOG TRANSFORMED (LOG<sub>10</sub>) DEATHS per MILLION (DpM) vs POPULATIONS' Vit. D LEVELS (nmol/L) |  |  |  |  |  |  |  |  |  |  |  |
|  |  | Data-Date | 12 Mar | 26 Mar | 8 Apr | 12 Apr | 26 Apr | 12 May | 26 May | 12 Jun | 26 Jun | 12 Jul | 26 Jul |
| Exponential Model<br>or Log<sub>10</sub><br>Transformed<br>Variable- Linear<br>Regression | Corr r(20)/R value |  | -0.3149 | -0.6024 | -0.6023 | -0.5899 | -0.5372 | -0.5156 | -0.5114 | -0.5085 | -0.5061 | -0.5084 | -0.5102 |
|  | R-square |  | 0.0991 | 0.3629 | 0.3627 | 0.3480 | 0.2886 | 0.2659 | 0.2615 | 0.2585 | 0.2562 | 0.2585 | 0.2603 |
|  | Adjusted R Square |  | 0.0491 | 0.3275 | 0.3273 | 0.3118 | 0.2491 | 0.2251 | 0.2205 | 0.2173 | 0.2148 | 0.2173 | 0.2192 |
|  | Standard Error |  | 1.3951 | 0.9007 | 0.5767 | 0.6042 | 0.5164 | 0.5120 | 0.5150 | 0.5197 | 0.5208 | 0.5204 | 0.5190 |
|  | F-value |  | 1.9807 | 10.2533 | 10.2452 | 9.6064 | 7.3023 | 6.5184 | 6.3749 | 6.2764 | 6.1988 | 6.2747 | 6.3330 |
|  | p-value |  | 0.1763 | 0.0049 | 0.0050 | 0.0062 | 0.0146 | 0.0200 | 0.0212 | 0.0221 | 0.0228 | 0.0221 | 0.0216 |

**Figure 1:**
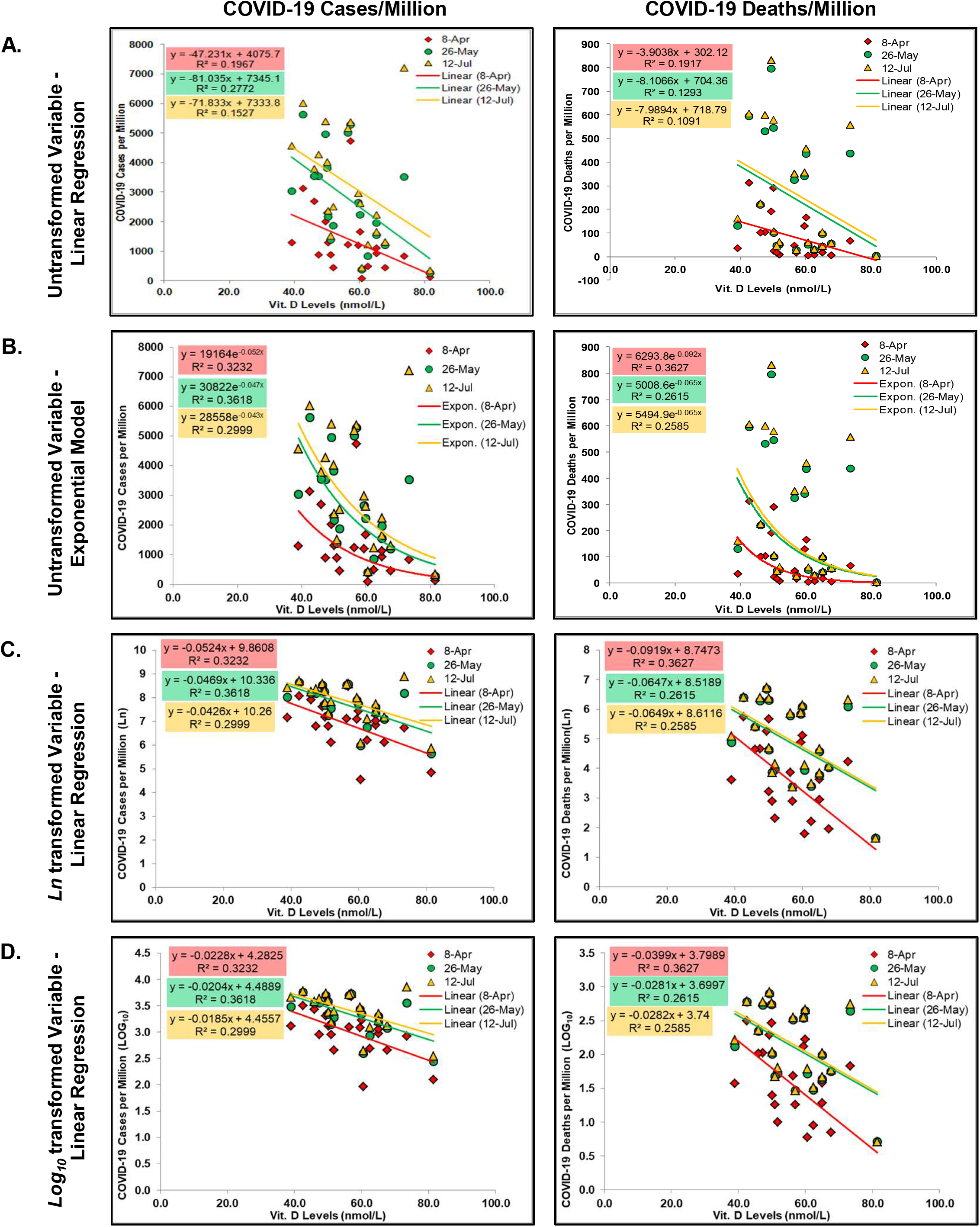
Time-series Regression analysis of COVID-19 impact and mean serum vitamin D levels in European populations. The vitamin D levels negatively correlated with the COVID-19 cases (left) and deaths per million populations (right) of the European countries during different time points of the study period. Representative analysis for 8 April, 26 May, and 12 July 2020 data is shown [See Table 1 for analysis of the period extending from 12 March (early pre-peak of infections) to 26 July (late post-peak of infections) 2020]. The correlation and regression estimates obtained on simple linear regression (A), improved on the application of exponential model (B) or linear regression model on transformed COVID-19 estimates of the population - *ln* (C) and *log*_*10*_(D). The transformation of data progressively suppressed the apparent variability/scatter in the data (compare 1C and 1D with 1B). The simple linear model was inadequate to support statistically significant (p-value<0.05) negative covariation of the COVID-19 incidences or mortality with vitamin D levels on 8^th^ April 2020 as well as at other time points analyzed (see Table 1).

The exponential correlative modeling of the data (Fig. 1 B) as suggested by us for the time period improved correlative predictive potential of populations vitamin D levels, *i*.*e*., *R*^*2*^ by 1.27-1.96 fold and *adjusted-R*^*2*^ by 1.33-2.47 fold for the COVID-19 cases per million, while the change in *R*^*2*^ by 1.81-2.67 and *adjusted-R*^*2*^ by 2.21-3.74 for deaths per million population upto the end of the current analysis period, *i*.*e*., 26 March (early stage) to 26 July 2020 (late infections post-peak), was observed. Its implementation over the simple linear regression was suggested by us for the exploratory statistical analysis of the time-series data preferably post-infections peak to arrive at more dependable estimates [7]. The simple linear model or regression of the log-transformed data (natural (*ln*) as well as *Log*_*10*_) for assessing potential predictive purpose (assuming cause and effect relationship is established in the future) displayed the same stable significant negative covariation (*p-*value<0.05) of the COVID-19 incidences and associated mortality/deaths in the per million population starting from 26 March (early stage) to 26 July 2020 (late infections post-peak) stage of the pandemic (Fig 1C and D and Table 1), with concomitant reduction in heteroscedasticity in the data set as expected (data not shown). Refer to Fig. 1 for the visual comparison of the data covariation (trend)/regression analysis performed, *i*.*e*., simple linear regression analysis, a more appropriate exponential curve fitting for the exploratory analysis, and the linear regression modeling/analysis of the data post *ln* and *Log*_*10*_ transformation of COVID-19 incidence and mortality per million populations. Note the transformation of data leads to visual suppression of underlying anomalies and any apparent heteroscedasticity that could have also indicated the presence of unidentified variable, with a progressively increased chance of missing out the important variable/anomalies that could have been discovered (compare Fig 1B with Fig. 1C and D).

## DISCUSSION

The race to discover potential protective variables for COVID-19 incidence and mortality is ongoing with a range of things being proposed to be correlated with COVID-19 impact [2-8] though not all necessarily having a potential protective role and without necessary rigor for such assertions. Based upon a correlation and regression analysis of a single day COVID-19 incidence and mortality data with serum vitamin D levels in populations [5], vitamin D had been proposed as a potential protective variable that may be explored further through dedicated studies. As indicated by us and others the one-day data analysis had weaknesses [7, 8] and it would have been better had the analysis been not constrained by commonly used *p*-value cutoffs and improper model application on the biological data set, and preferably estimating the correlation post-peak of infections to reduce the effect of potential confounders of data reporting delay in infection and adverse outcome, and analyzing the correlation or regression at multiple points to arrive at potential dependable estimates.

We now have COVID-19 data for the European countries for a larger time frame that by 26 July 2020 included a total of 1,829,634 cases and 179135 deaths accounting for the worldwide 11.11% cases and 27.45% deaths. The reanalysis of the data for an extended period indicates the potential problem of the analysis and model presented by Ilie et al. [5] which was previously predicted/indicated by us and others [7,8]. The simple linear regression modeling of the covariation of vitamin D levels with COVID-19 cases per million as suggested by the authors did not indicate a statistically significant correlation (p-value≥0.05) upto 8 Apr, which became statistically significant (at p-value cut off <0.05) by 12 April and stayed that way till 12 Jun then again become insignificant and remained so till the end of the analysis period, *i*.*e*., 26 July 2020. The correlation between deaths per million and vitamin D levels *never* ever reached the commonly used statistical significance level suggested by Ilie et al. during the whole study period starting from 12 March to 26 July including that on the 8^th^ April 2020 when it was most closer to the significant p-value cutoff of 0.05 [*r*(20) - 0.4378, *p*-value 0.0535]. In the study period, the *p-*value had actually progressively increased from 0.0535 on 8^th^ April to 0.1565 on 26 July 2020. Thus, the reanalysis of indicated 8^th^ April data as well as the time-series data of the period extending upto 26 July 2020 using previously suggested methodology to model or evaluate the vitamin D levels and COVID-19 impact on the populations, could not endorse a correlational potentially protective conclusions drawn about the populations mean serum vitamin D levels.

However, exponential modeling of the covariation (or the linear regression analysis of vitamin D levels and *ln* / *log*_*10*_ transformed COVID-data) improved the correlation as well as correlative predictive potential over the simple linear regression analysis upto the end of the current analysis period, *i*.*e*., 26 March to 26 July 2020. For the COVID-19 cases per million population the *R*^*2*^ and *adjusted-R*^*2*^ increased by 1.27 - 1.96 and 1.33 - 2.47 fold, respectively while for deaths per million population, *R*^*2*^ and *adjusted-R*^*2*^ increased by 1.81-2.67 fold and 2.21-3.74 fold, respectively. As apparent in table 1 the standard error term, F-value and p-values combined show a drastic transition between 12 March (early stage of COVID-19 pandemic) to 8-12 April 2020 then stabilizes and remains so. More importantly, the analyzed parameters (correlation, *R*^*2*^, adjusted-*R*^*2*^, *p*-value) remain more stable post-infections-peak (12 May 2020) as predicted and suggested previously by us, suggesting the potential existence of a cause and effect relationship between Vitamin D levels and COVID-19 impact that may be tested in dedicated future studies.

However, some of the limitations about the available data and hence the analysis presented must be borne in mind while designing experiments/trials to explore the protective potential of serum vitamin D in COVID-19. The presence of data on the overall deficiency or sufficiency variation in vitamin D levels of the affected populations or the individuals who got affected or had an adverse outcome in the current pandemic would have significantly improved the confidence of our analysis. Robustness of the analysis could have been further improved if the data of mean serum vitamin D levels of other European countries that have had experienced the moderate impact of COVID-19, would have been available and included in the analysis - as the countries currently included in the analysis seemingly are from affected extremes (severe and low) accounting for 11.11% of the cases but 27.45% deaths worldwide opposed to 18.34% cases and 31.73% deaths accounted by European countries combined together on 26 July 2020 [1], so more prone to biases due to relative disparate data sets [9]. Nevertheless, the potential protective impact of higher (or lower) vitamin D levels on COVID-19 incidence and adverse outcome possibility could be ascertained through controlled trials to ascertain its actual effect so as to limit any adverse impact due to prevailing vitamin D levels or alternatively that may be resulting from over or under supplementation of vitamin D with or without a medical prescription in the populations.

The analysis as such presented, also highlights the pitfalls of our fascination with p-value cutoffs, potential data dredging knowingly or unknowingly, and the need for more rigor exercised by authors, reviewers, and editors of the journal alike in the current testing times [Refs in 8]. The ongoing urge or fascination to go on to fit the data to unprecedented level in literature and the apparent data dredging seen during the current COVID-19 pandemic has to be balanced. It has been observed besides the necessary exploratory statistical analysis of the existing data to find any variable that could be potentially protective or be making individuals vulnerable that could be evaluated clinically or through controlled trials, more efforts appear to be invested in making the correlations look significant by inclusion of the unequal data points, the inclusion of variables logically not seemed to have cause and effect relationship, and the transformation of the variables to derive higher association/covariation/correlation and the highly significant ‘lower unrealistic p-value’ without any metrics for size effect wherever possible. We do agree that statistical modeling showing highly correlated variables should be evaluated with caution and the journals/editors may take a lead in informing/educating the audience and reviewers alike as had been advocated in the recent past [References in 8]. In the same vein as indicated by Maruotti *et al*., we have presented the perils of the one-point data analysis in the exploratory epidemiological study and indicate how the data analysis methodology employed by Ilie *et al*. may have been inadequate to derive the conclusions. We would submit that the senior editors of esteemed journals could take a more precautionary approach in both educating the junior editors and reviewers about the perils of statistical analysis and one point cross-sectional analysis, the fascination of unrealistic p-values that may make something appear significant when they are not as p-values drastically change with the number of data points in a small data set as well as the inclusion of selective disparate data sets. Equally important would be to avoid visual depiction/presentation of only transformed data as much as possible in the exploratory analysis so as to allow discovery of other potential variables that may otherwise remain hidden in the plain sight on generally employed data transformations such as *ln* / *log*_*10*_, square root, etc. The least transformed (minimum necessary) data presentation could allow discovery of potential protective variables not necessarily by the authors of the study who may have already missed it anyway or had reasons to believe it was not important, but by onlookers who may have access to knowledge of other variables. For instance in the data set presented the population of Sweden (mean vitamin D level 73.5nmol/L) that was apparently more protected from COVID-19 infection on 8 April 2020 as compared to many countries, *e*.*g*., Spain, Iceland, Italy, Switzerland, Denmark, Norway had surpassed all nations by 12 July 2020, potentially due to a variable only applicable to Sweden. Nonetheless, the stark outlier could be more easily identified in the untransformed data presentation by someone who may know that Sweden as a nation among all the countries analyzed had one of the lowest stringent measures in place to prevent the spread of COVID-19, so increasing the chance of further refinement of the analysis presented. Similarly, other readers/scientists who may be short on time could also contribute to the identification of other variables which may be helpful in the long run to find out a solution to the problem in hand. So, in our view, for the exploratory statistical analysis, the data may be presented with the least transformations employed to increase the chances of the discovery of new variables. The heteroscedasticity may be allowed to stay in plain sight, rather modeling parameters could be changed to show the potential relationship and in no way the exploratory regression analysis be construed as a one depicting cause and effect relationship. We like to add, for all the exploratory analyses performed the authors may also proactively reassert at quite a few places that indicate the analysis to be exploratory in nature or some consensus among the scientific fraternity could be arrived to clearly label such studies.

In conclusion, our time-series cross-sectional epidemiological exploratory study using European countries COVID-19 incidence and mortality data starting from 26 March to 26 July 2020 indicates consistently negative (or potentially protective) covariation with serum vitamin D levels, and suggest vitamin D as one of the potential candidates for evaluation by dedicated control trial studies using matched (i.e., age, comorbidities, sex, genetic background, disease severity, etc) control and test groups to once and for all establish the role of vitamin D levels on COVID-19 incidences or the adverse outcome if any whether negative or positive. The controlled trials initiated in different countries for evaluating the link between vitamin D levels and COVID-19 outcomes, e.g., Argentina (NCT04411446), France (NCT04344041), Iran (IRCT20200324046850N1), Spain (NCT04334005), could provide the necessary evidence for the same [10].

## Data Availability

Included in the manuscript and available at indicated sources.

## Ethical statement

The study is compliant with the ethical standards. Considering the design of the study no human or animal rights were infringed upon.

## Informed consent

Considering the design of the study no informed consent was necessary.

---

**Supplementary Table 1. Sheet 1, labeled ‘12 Mar - 26Jul CpM DpM & Vit D Lev’:** Estimates of Vitamin D levels (nmol/L) and COVID-19 incidence and mortality (per million) during study period 12 March to 26 July 2020 [1,5], along with basic statistical estimates; **Sheet 2, labeled ‘Regression Analysis Table’** same as Table 1 coming from **Sheet 3, labeled ‘Regression Analysis Results’:** Raw data of regression analysis

